# A model of endemic coronavirus infections

**DOI:** 10.1101/2020.11.08.20227975

**Authors:** David S. Huen

## Abstract

This work proposes that epidemiological features of both endemic coronaviruses and the recent highly pathogenic outbreak coronaviruses can be combined within an integrated framework. In this framework, mortality amongst those infected for the first time is mostly amongst the old but survivors acquire fatal infection immunity (FII). Subjects with FII can subsequently be infected and infect others without suffering significant mortality. Under these conditions, coronaviruses induce endemic infections that elicit FII in individuals during childhood when the risk of mortality is low and maintain it throughout their lifetime, thereby protecting the population against the worst effects of infection.

A multi-compartment ODE model was constructed to explore the implications of this proposal on the evolution of a zoonosis sharing properties of both SARS-CoV-2 and endemic coronaviruses. The results show that mortality has two components, the first incurred during transition to endemicity and the other is exacted on a continuing basis. The relative contribution of each depends on the longevity of the FII state. In particular, a one-time vaccination of the older subpopulation is sufficient to reduce total mortality if FII is long-lived. The effect of a regular vaccination was also examined when FII was shorter lived. Herd immunity was not achieved.

The validity of this proposal with regard to Covid-19 depends on whether reinfection with SARS-CoV-2 behaves in the manner expected of FII. If it does, then certain considerations apply to how Covid-19 is to be managed and how vaccine choice could influence that.

## Introduction

Human endemic coronaviruses (ECoV) comprise four coronaviruses (229E, HKU1, NL63 and OC43) of minor clinical importance (reviewed in Ogimi *et al*, 2020). They are responsible for a range of upper respiratory tract infections, infecting most humans from an early age and throughout life (Zhou *et al*, 2013; Ogimi *et al*, 2020; Dijkman *et al*, 2012; Huynh *et al*, 2012). Infections can be asymptomatic (Prill *et al*, 2012; Walsh *et al*, 2013; Morikawa *et al*, 2015). Phylogenetic evidence suggests zoonotic origins for NL63 (from bats, 1190-1449; Fouchier *et al* 2004; Huynh *et al*, 2012), 229E (from bats, 1686-1890; Pfefferle *et al*, 2009) and OC43 (from cows, 1890s; Vijgen *et al* 2005). While their clinical presentation is similar, they differ in receptor use, with OC43 and HKU1 targeting 9-O-acetylated sialic acid moieties on surface proteins (Hulswit *et al*, 2019), 229E targeting aminopeptidase N and NL63 targeting ACE2 (Fouchier *et al*, 2004).

The ubiquity of ECoV infections suggests ECoV regularly reinfect humans and antibody titre evidence exists demonstrating this (Eldridge *et al*, 2020). Being of minor clinical significance, the immunology of ECoV infection has been neglected. While antibody titres decline soon after infection, reinfection can and does occur while significant serum antibody titres still persist (Callow *et al*, 2010; Eldridge *et al*, 2020). Mucosal antibody titres are likely to be more important than serum antibody titres in preventing reinfection: while the latter were readily detected in older adults, the mucosal wash IgA was only detected in a minority (Gorse *et al*, 2010). It is not known how ECoV infections are immunologically resolved and while innate and cell-mediated immunity can be expected to play key roles, their role has yet to be investigated.

Understandably, the emergent highly pathogenic coronaviruses responsible for zoonoses over the last two decades have received more attention. As with ECoVs, SARS antibody titres declined after infection (Ho *et al*, 2005; Mo *et al*, 2006). Covid-19 is too new a disease to provide a long observational baseline but the balance of evidence suggests declining serum antibody titres too (Ward *et al*, 2020; Long *et al*, 2020; Seow *et al*, 2020). In contrast, T cell immunity appears to be sustained long term, with responses detected up to 17 years later with SARS (Ng *et al*, 2016; Le Bert *et al*, 2020). Strong T cell responses were also elicited by SARS-CoV-2 (hereafter referred to as SARS-2), the causative agent of Covid-19, in both mild and severe cases (Le Bert *et al*, 2020; Braun *et al*, 2020; Peng *et al*, 2020; Sekine *et al*, 2020) and the immunological differences between these outcomes is now emerging (Laing *et al*, 2020). Surprisingly, T cell responses to SARS-2 targets were discovered in uninfected controls in some of these studies but not others (Le Bert *et al*, 2020; Braun *et al*, 2020; Nelde *et al*, 2020; Bacher *et al*, 2020; Peng *et al*, 2020). These T cells have been attributed to cross-reactive T cells and the conflicting results to technical differences between studies. However, those identified in one study have been shown to be of low avidity and may the authors propose they are related to age and detrimental to disease outcome (Nelde *et al*, 2020). Even severe coronavirus infections like SARS and Covid-19 can have an asymptomatic component (Lee *et al* 2003; Wilder-Smith *et al* 2005; Kronbichler *et al*, 2020).

A distinctive feature of SARS, MERS and Covid-19 is that mortality is strongly linked to old age and co-morbidities (Chan-Yeung and Xu, 2003; Mizumoto *et al*, 2015; Park *et al*, 2018; Ghisolfi *et al*, 2020). The young are largely spared when infected. Recent work demonstrated that defects in the the innate immunity pathway were associated with severe Covid-19 indicating its importance in controlling SARS-2 (Zhang *et al*, 2020) and declining innate immunity is a prominent feature of the aging immune system (Fulop *et al*, 2013; Montgomery and Shaw, 2015; Boe *et al*, 2017).

I suggest that the characteristics of ECoV and the more recent highly pathogenic viruses can be married within a model based on:-

- A S(usceptible) state where mortality during infection increases with age: non-immune subjects will be in this state.
- A V(iable) infectible state with negligible mortality. The V state is persistent, possibly lifelong.
- On surviving an infection, all subjects are briefly resistant to reinfection then enter the V state.

The existence of the S state cannot be readily demonstrated with ECoVs as they will have infected the entire population during their youth when S state mortality is expected to be negligible. Conversely, the mortality profiles of SARS and Covid-19 are very much what is expected of the S state. Also, the recent coronavirus zoonoses show varying mortality with Covid-19 < SARS < MERS. ECoV are also capable of fatal lower respiratory tract infections so they cannot be precluded *a priori* from also possessing age-dependent S state mortality profiles (Gorini da Veiga, *et al*, 2020). The OC43 zoonosis has been suggested as the true cause of the Russian flu pandemic of the 1890s (Vijgen *et al*, 2005).

In contrast, ECoVs clearly show V state behaviour with frequent reinfections separated by short periods of resistance to reinfection. The severity of MERS and SARS drove strenous efforts that eradicated those viruses before reinfection was observed. Covid-19 is too new to have observed significant levels of reinfection. Of the five known cases, none have been fatal. SARS immunology showed antibody responses to be short-lived but T cell responses were maintained over many years. This would be consistent with the V state characteristics if antibody responses correlated with protection from reinfection and T cell responses with protection from fatal infection.

Within this model, ECoV infections are endemic and maintained in the community by the absence of long-term immunity to reinfection. Long term herd immunity cannot be attained under these conditions. Humans are first infected and immunologically primed as children when they are least likely to exhibit fatal outcomes. If fatal infection immunity is lifelong, circulating infections are responsible for its establishment amongst neonates. If not, it plays an additional role in maintaining FII throughout the population such that completely losing immunity and returning to the S state is uncommon and fatal infection is therefore an rare outcome. It is noted that the recent coronavirus zoonoses originated from bats and they have a dense social structure ideally suited to maintaining high rates of infection throughout their lifespan. It is not therefore necessary to invoke unique physiological characteristics to explain why they can sustain high levels of coronavirus infections without fatality if bat viruses have ECoV characteristics.

Again, this proposal predicts that when a new ECoV zoonosis occurs, fatalities only occur amongst the non-immune part of population and then, mostly amongst the old. Thereafter, a lower rate of mortality is experienced, that rate being dependent on how fast FII is lost. With lifelong FII, the maximum death toll that can be suffered from such a zoonosis is that obtained by applying the age- and sex-specific infection fatality rate (IFR) profile to the demographic profile of that population. For example, for the UK, IFR estimates range up to 0.9% and a maximum potential mortality is 600 thousand persons (Ward *et al*, 2020). By way of comparison, there are around 600 thousand deaths annually in the UK so if Covid-19 induces lifelong FII, its *theoretical* worst case outcome is equivalent to the deaths that can be expected in a year although that would certainly exceeded in practice when critical care facilities are overwhelmed. Since Covid-19 deaths are mainly amongst the old, this will also result in depressed excess deaths in the next few years. Fatalities from ECoV zoonosis are effectively self-limited if FII is long-lasting.

There are also implications for control measures. Since circulating infection is a key protective aspect of ECoV infection, control measures need to lower mortality without hindering the transition to endemicity.

This study modelled the behaviour of ECoV infections and interventions in the transition towards endemicity.

### Model Structure

The model is a multi-compartment ODE model (figure 1). The S state has already been described. The R(ecovered) state represents a short period of immunity to reinfection enjoyed after recovering from an infection. Two states of infection are specified. The potentially F(atal) state represents the infection of a naive subject and it resolves by transiting to either the R state or the D(ead) state, the partition between the two being determined by an age- and sex-dependent infection fatality rate (IFR). The alternative infected Y state represents reinfection of an already immune host and only resolves non-fatally to the R state.

**Figure 1:**
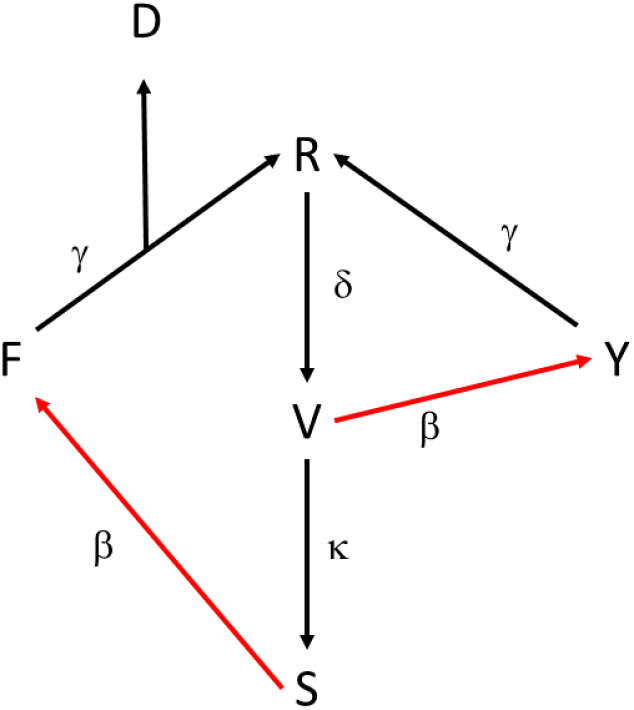
Structure of ODE model.

The V state represents those possessing FII, who are infectible but will not be killed by the virus. If FII is not lifelong, subjects exit the V state to return to the S state.

Each state has an internal age profile of 90 year-wide bins for each of the sexes. Aging was modelled in all states that represent living subjects to permit long-term evolution of the model.

Rate coefficients are indicated next to the edge that represents that state transition in figure 1.

Further details of the model are available in the Material and Methods.

## Results

### Transition to endemicity

The implications of these ECoV chracteristics was investigated in an contact matrix-based ODE model. A population of 1 million individuals was simulated, setting *R*_0_ = 2.5 with recovery from infection as a first-order process with a 6 day half-life. Resistance to reinfection was lost with a 1 year half-life to model the interval between infections experienced with ECoVs. The simulations were initially performed assuming lifelong FII (*κ* = 0) and varied later.

Two populations were initially modelled with England+Wales as the base case and Singapore as the alternate case, using age, sex and mortality profiles and contact matrices specific to each. As the long-term behaviour of the endemic was of interest, the model also incorporated aging of the population. The birth rate was continuously adjusted to maintain a constant population. The two populations represent economically well-developed communities at different stages in the demographic transition with Singapore being at an earlier stage. Consequently, its population structure is younger than that expected from its age- and sex-specific mortality rates and its birth rates in the population rose later in simulation to balance increased deaths from aging. The infection fatality ratio profile used in the simulation was the High Income Country set for Covid-19 reported by Ghisolfi *et al*, 2020.

All simulations were seeded with an infected, 20 year old male in the F state.

The base case examined the natural evolution of a zoonosis over a 50 year span where the seeded population was initially in the non-immune S state (figure 2(a)).

**Figure 2:**
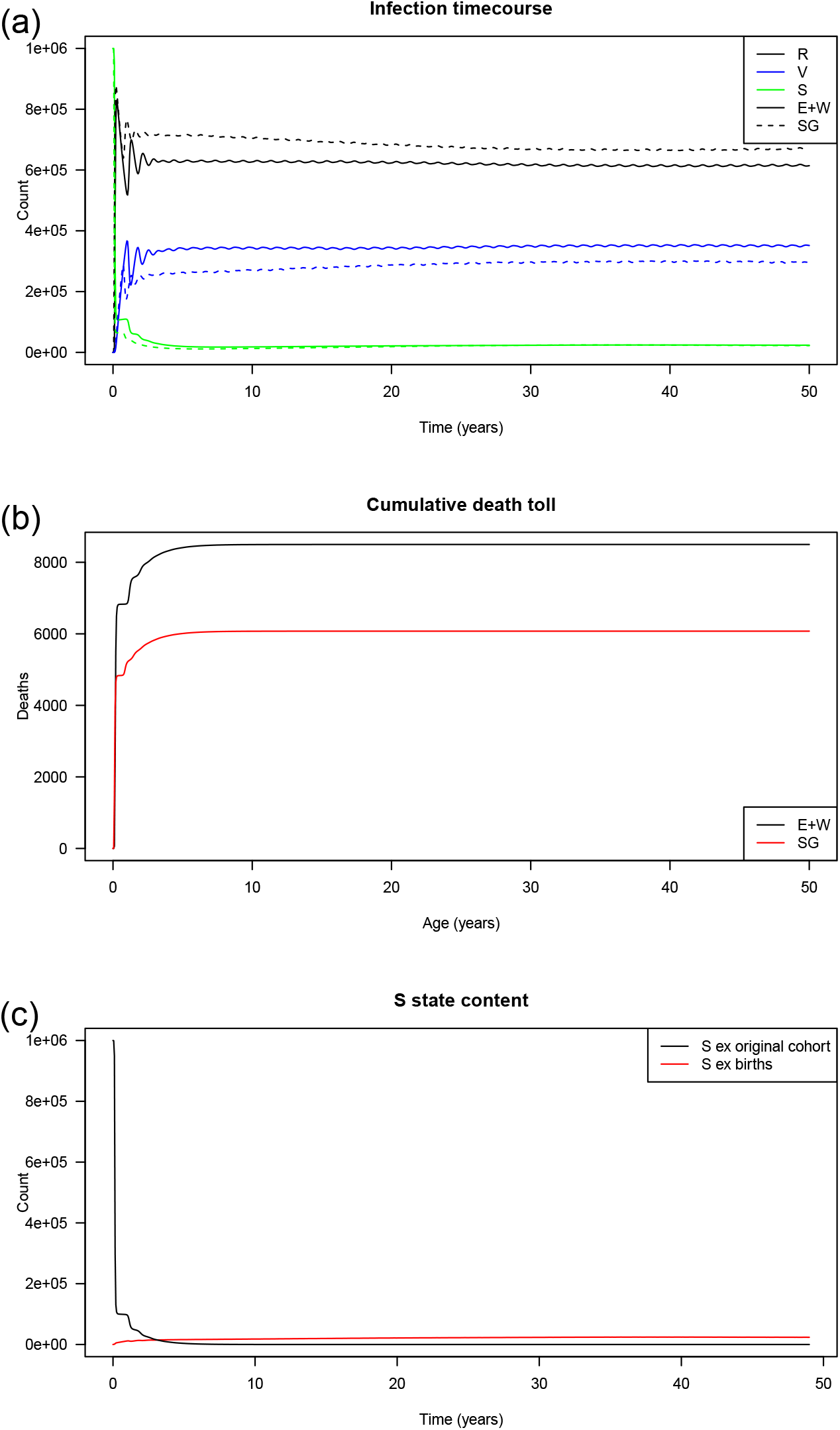
Evolution of outbreak simulations. (a) The occupancy of R, V and S states over a 50 year period are displayed for both the E+W (solid line) and SG (dashed line) populations. (b) Cumulative deaths are shown for the E+W (black) and SG (red) populations. (c) Number of subjects in S state, differentiated on whether they were from the original cohort (black) or from new births (red).

Both populations showed qualitatively similar behaviour (figure 2).

It can expected that without a lifelong uninfectible state, infection would spread throughout the population and the surviving population will be refractory to fatal infection thereafter, new births being immunized by circulating infection. However, following an initially rapid spread, progress thereafter was slower and around a half decade was required to converge towards the same equilibrium. This can be attributed to a third of the population being in the short-term uninfectible R state and subsequently temporarily hindering the spread of infection. This is also observed with deaths continuing to rise for many years (figure 2(b)). Deaths were higher in England+Wales than Singapore, as expected since the former is an older population (figure 2(b)).

It was noted that the S state did not decline to zero with time and that appeared to contradict the earlier claim of complete infection (figure 2(a)). To explore this further, a modified dual model was built in which every state was duplicated and the paired submodels were linked via infection. A cohort could therefore be transferred to and segregated within the secondary submodel and its fate tracked through an entire lifetime. When births and the initial cohort were segregated, it can be seen that the subjects in S state initially were completely depleted by infection and the subsequent occupants of the S state were derived from new births (figure 2(c)).

From the foregoing, fatalities would occur only amongst the initial cohort with lifelong FII. A crude estimate of that mortality can be obtained by applying the age- and sex-specific IFR to that cohort assuming that they would be promptly infected. Using the segregated model, disease mortality of cohorts of each 10 year age group were tracked across their entire lifetimes and compared against the IFR-derived estimates (tables 1 and 2). The results closely matched expectations at most age categories.

**Table 1:**
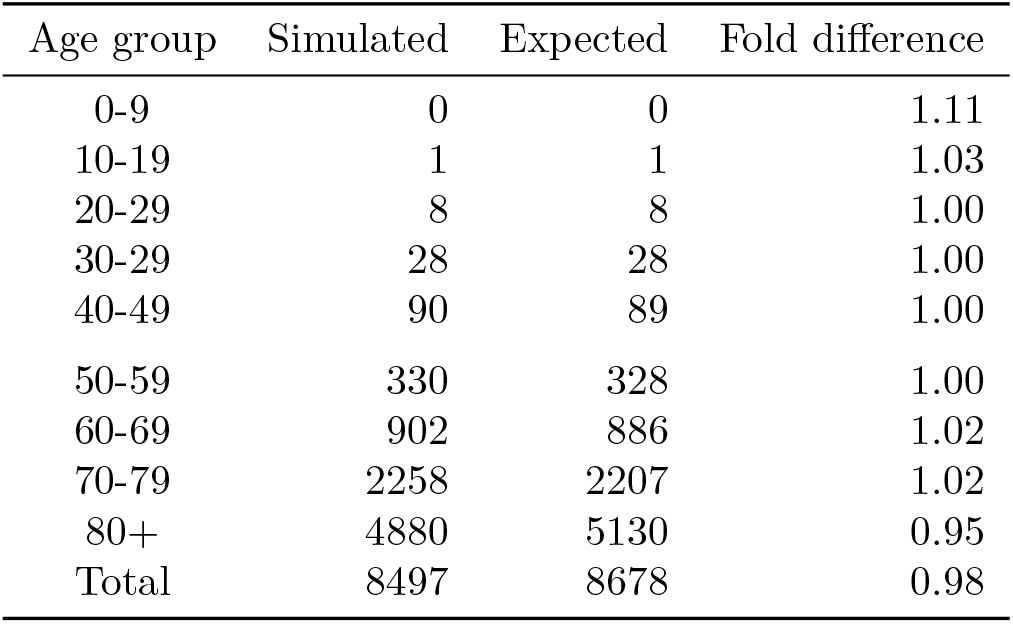
Comparison of deaths obtained from simulation against that calculated from IFR - England+Wales.

| Age group | Simulated | Expected | Fold difference |
| --- | --- | --- | --- |
| 0-9 | 0 | 0 | 1.11 |
| 10-19 | 1 | 1 | 1.03 |
| 20-29 | 8 | 8 | 1.00 |
| 30-29 | 28 | 28 | 1.00 |
| 40-49 | 90 | 89 | 1.00 |
| 50-59 | 330 | 328 | 1.00 |
| 60-69 | 902 | 886 | 1.02 |
| 70-79 | 2258 | 2207 | 1.02 |
| 80+ | 4880 | 5130 | 0.95 |
| Total | 8497 | 8678 | 0.98 |

**Table 2:**
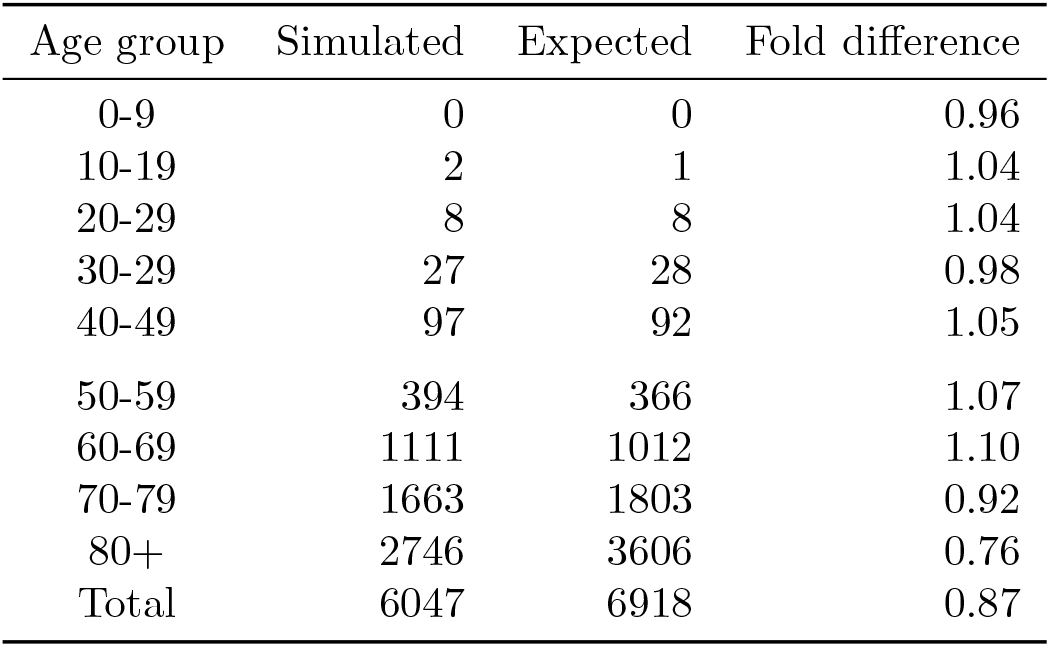
Comparison of deaths obtained from simulation against that calculated from IFR - Singapore.

The median time to fatal infection (MTFI) and lifetime IFR (LIFR) were also estimated for the base case (tables 3 and 4). With lifelong FII, the majority of fatalities occur in the first wave of infections (table 3, 2nd column). With finite FII halflives, subjects occasionally return to the S state over their remaining years, where they were exposed to a rising risk of fatal infection. The MTFI for age groups indicate that the majority of virus-caused deaths occurred after they entered their late 70s (table 3). The LIFR also show virus deaths rising with both initial age and *κ* (i.e. shorter FII halflives). The former can be explained by older age groups having higher risks of mortality during their initial, unprotected infection. The young will have FII by being infected while their risk of FII was low. The risk of loss of FII increases with *κ*, eventually exposing a fraction of those cohorts to virus mortality in their old age.

**Table 3:** Median time to fatal infection (MTFI) for population at start of outbreak in the England+Wales scenario

| Age group | Lifelong FII | $\kappa = 0.025$ | $\kappa = 0.05$ | $\kappa = 0.10$ | $\kappa = 0.20$ |
| --- | --- | --- | --- | --- | --- |
| 0-9 | 0.2 | 80.9 | 80.8 | 80.6 | 80.2 |
| 10-19 | 0.2 | 71.1 | 71.0 | 70.8 | 70.4 |
| 20-29 | 0.1 | 60.9 | 60.8 | 60.6 | 60.2 |
| 30-29 | 0.1 | 50.8 | 50.8 | 50.7 | 50.3 |
| 40-49 | 0.1 | 40.2 | 40.4 | 40.4 | 40.1 |
| 50-59 | 0.1 | 28.7 | 29.8 | 30.2 | 30.1 |
| 60-69 | 0.2 | 11.1 | 17.1 | 19.1 | 19.8 |
| 70-79 | 0.2 | 0.2 | 1.7 | 6.7 | 9.4 |
| 80+ | 0.2 | 0.2 | 0.2 | 0.2 | 0.3 |

**Table 4:** Remaining lifetime IFR (%) for population at start of outbreak in the England+Wales scenario

| Age group | Lifelong FII | $\kappa = 0.025$ | $\kappa = 0.05$ | $\kappa = 0.10$ | $\kappa = 0.20$ |
| --- | --- | --- | --- | --- | --- |
| 0-9 | 0.0% | 1.1% | 2.1% | 3.9% | 6.7% |
| 10-19 | 0.0% | 1.1% | 2.1% | 3.9% | 6.7% |
| 20-29 | 0.0% | 1.1% | 2.1% | 3.9% | 6.7% |
| 30-29 | 0.0% | 1.1% | 2.1% | 3.9% | 6.7% |
| 40-49 | 0.1% | 1.2% | 2.2% | 4.0% | 6.8% |
| 50-59 | 0.2% | 1.4% | 2.4% | 4.2% | 7.1% |
| 60-69 | 0.9% | 2.0% | 3.0% | 4.9% | 7.8% |
| 70-79 | 2.7% | 3.8% | 4.8% | 6.5% | 9.3% |
| 80+ | 9.7% | 10.4% | 11.1% | 12.3% | 14.3% |

This model has a *force of infection* (FOI) that is common to to all subjects within an age-group, that is, the per-person rate of infection is the same for all within that age group. The FOI for the *a*th age group is described by

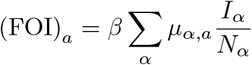

where *I*_*α*_ is the number of infected and *N*_*α*_, the size of the *α*th age group in the population. *µ*_*α,a*_ are terms linking the *a*th and *α*th age groups in the contact matrix.

The infection rate for a subpopulation at risk is then just the product of the size of that subpopulation with the appropriate FOI for that age-group. Note that the FOI underwent sharp transients early in the infection that eventually decayed to their equilibrium values (figure 3(a)). The value of FOI can be considered as the average number of infections per person per unit time so its reciprocal is roughly the time between infections for that age group, neglecting time spent in the *R* state. It was noted that the FOI was lower for both the very young and the very old (figure 3(b)). For the England+Wales model, the 75+ year year old age group had an FOI of 0.67 which corresponds to an infection every 1.5 years although the average of a year spent in the *R* state would increase that. This should be adequate to maintain a fair degree of FII at lower *κ* values. The combination of reduced FOI and increased mortality from other sources also negated some of the effect of increased IFR amongst the 80+ age group.

**Figure 3:**
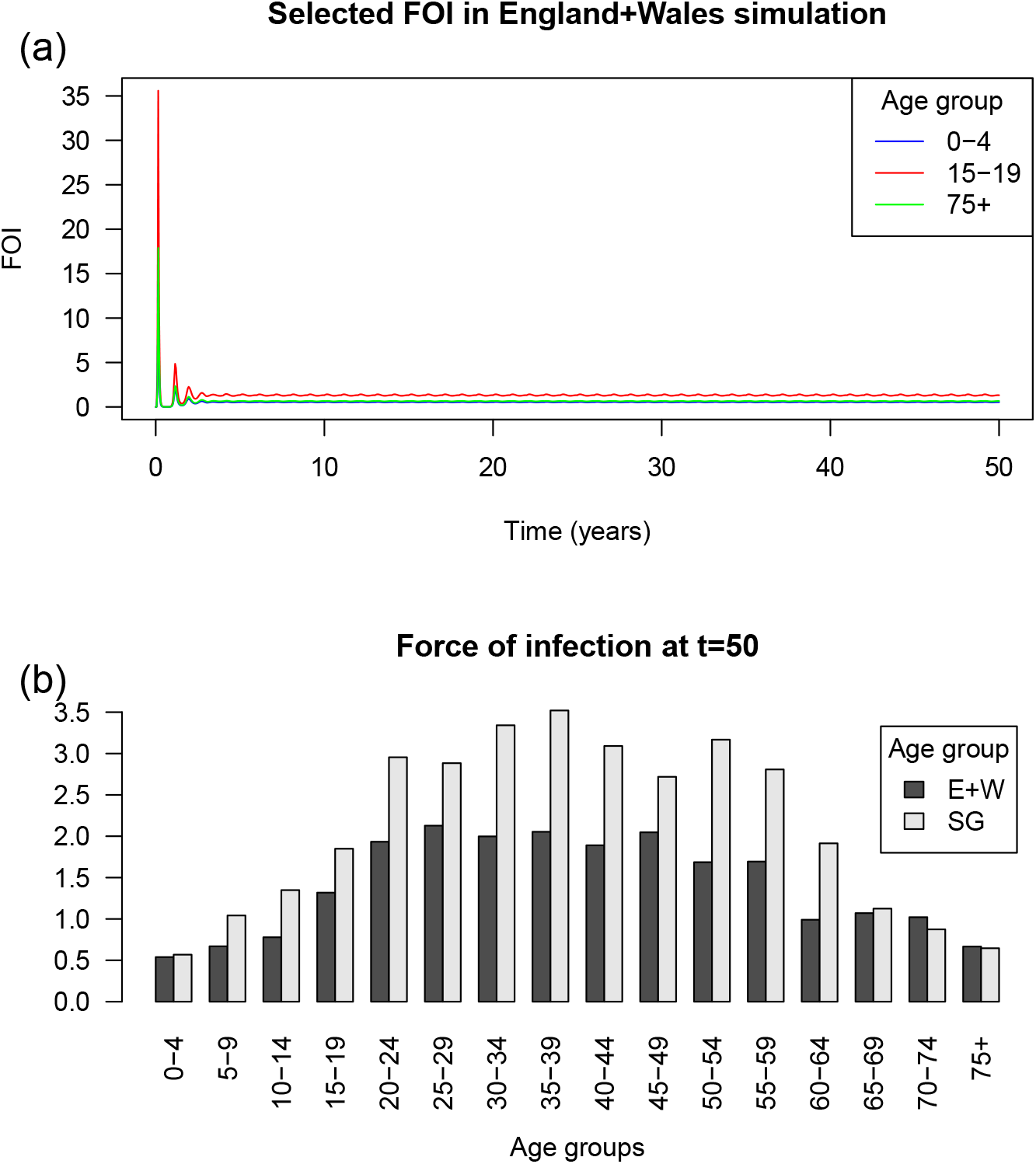
Force of infection. (a) FOI values for 0-4, 15-19 and 75+ age groups are shown for England+Wales simulation. (b) Distribution of FOI at near-equilibrium values are shown for E+W and Singapore simulations.

### Limited duration immunity

The foregoing analysis assumed lifelong immunity to fatal reinfection. The effects of this immunity being long-lasting but not lifelong is now examined. A particular concern is that the reinfection could be too infrequent to maintain fatal infection immunity (FII).

The loss of FII is represented in the model by a V → S first-order decay that is parameterized by *κ*. Values ranging from *κ* between 0.025-0.2 were examined, which correspond to V decay half-lives of 27.7 years down to 3.5 years. Non-zero values of *κ* had the effect of increasing the occupancy of the S state associated with mortality and decreasing that in the V state which is not subject to fatal infection figure 4(a). However, while the V → S transfer was not age-specific, age-specific variations in contacts resulted in biased age distributions for the S and V states. The mid-range age groups were reinfected and retained in states within the non-fatal infection loop (R,V and Y) by their higher numbers of contacts while the young and old were preferentially lost (figures 4(b); see figure legend for explanation of spike at age 50). The old were swept into the S state with concomitant loss of FII wherein they were preferentially retained for the same reason (figure 4(c)). The young started in the S state but were steadily removed by infection from the S state to the V state (figure 4(c)). In contrast, the limited residual life expectancy of the old resulted in other causes of mortality claiming a proportion of those in the S state before they could succumb to the virus.

**Figure 4:**
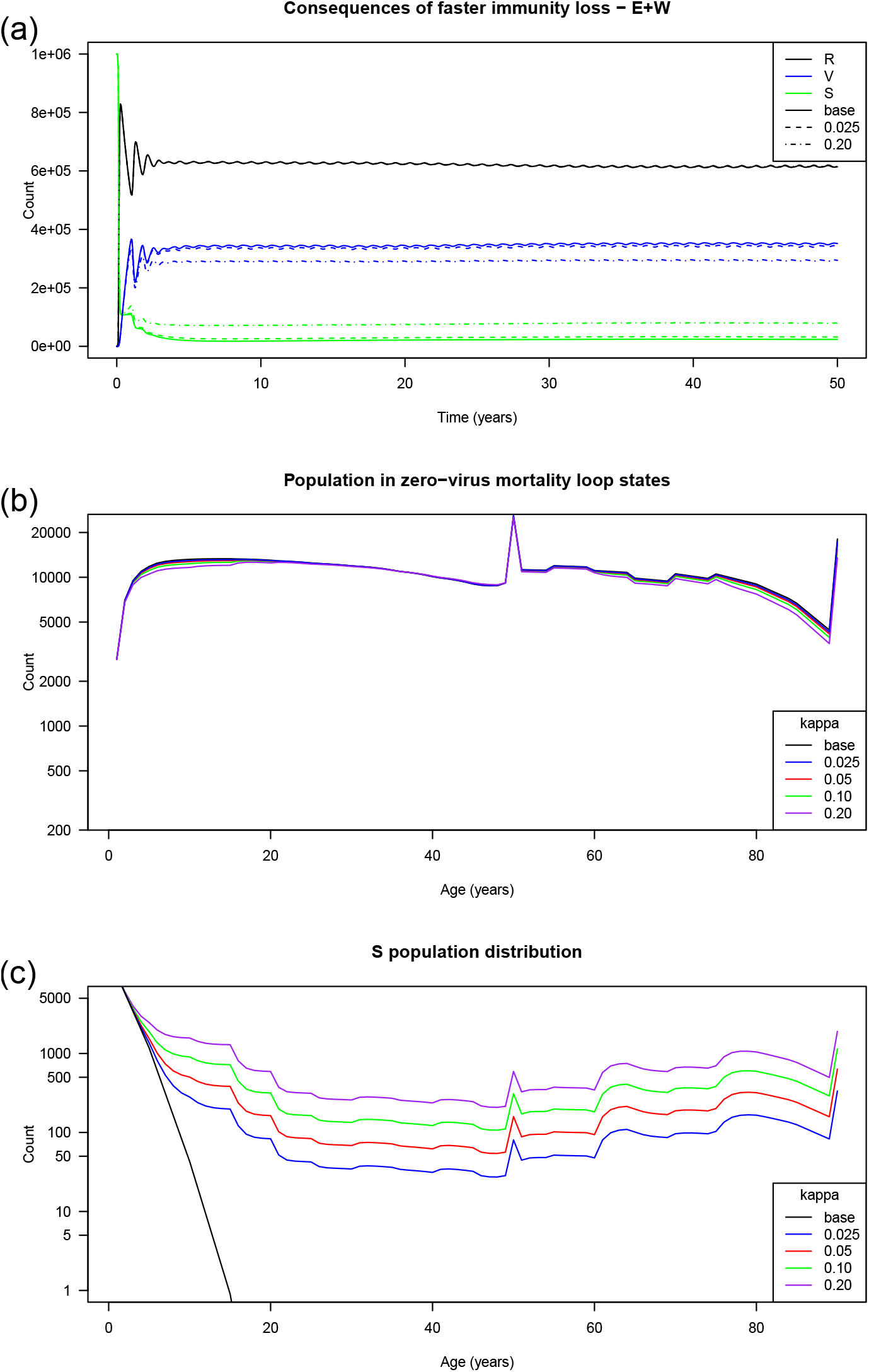
Effect of differing rates of immunity loss. (a) time course of S, V and R state occupancy with lifelong FII and a low (*κ* = 0.025) and high (*κ* = 0.2) rates of loss. (b) Age distribution of V state at varying rates of immunity loss. (c) Age distribution of S state at varying rates of immunity loss. The large spike around age 50 corresponded to births injected by the model to balance the high number dying early in the outbreak. Note that the counts at the younger ages were swelled by new births. The final bin accumulated all subjects older than 89.

Mortality with limited term FII can be characterised by an initial, one-off *transitional mortality* (figure 5(a)) followed by a transition to a long term, equilibrium endemic mortality rate that was dependent on *κ* (figure 5(b)). Whereas the endemic mortality rate was zero with lifelong FII, it rose with increasing values of *κ* as decay of FII exposed increasing numbers of subjects to fatal reinfection.

**Figure 5:**
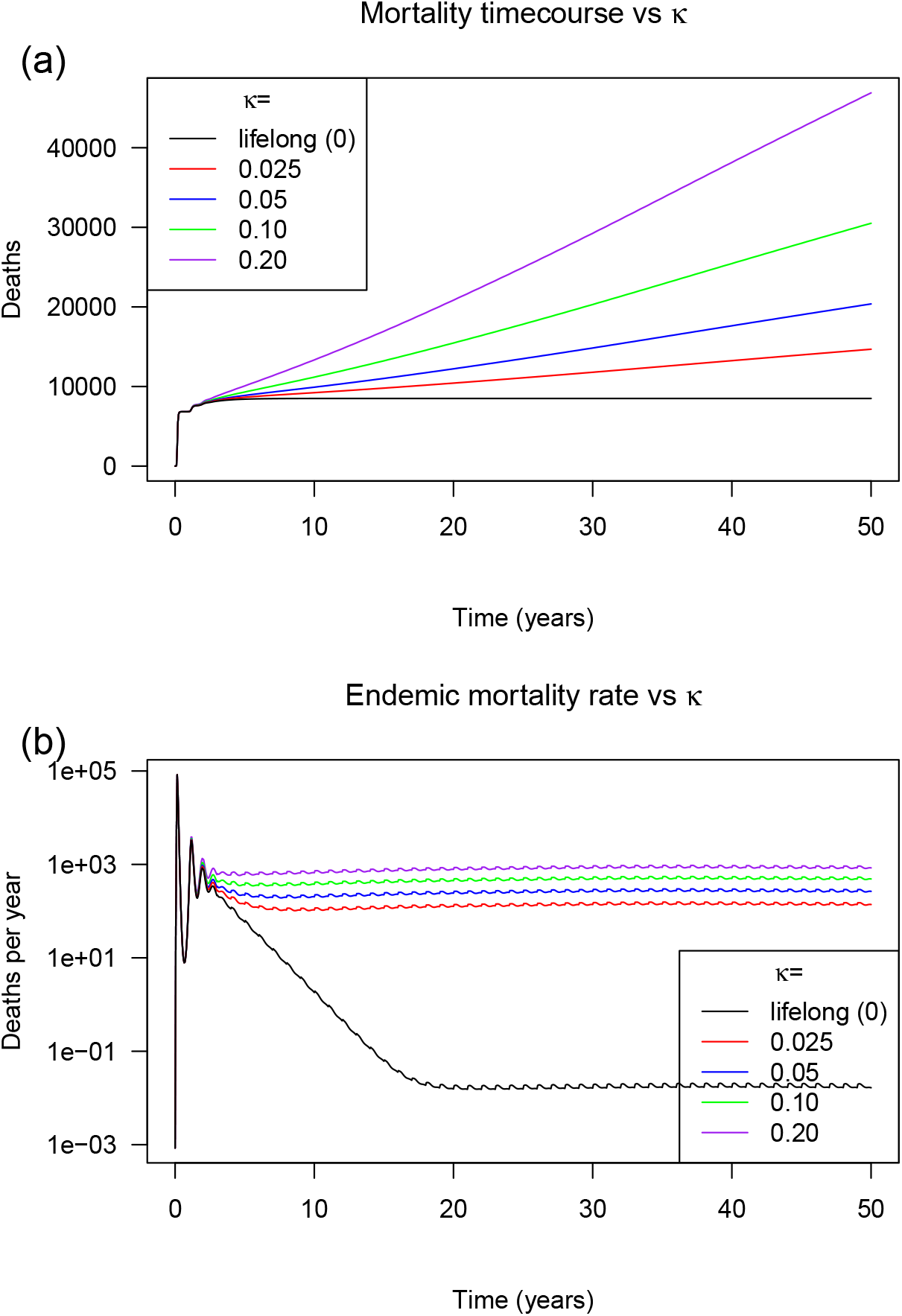
Mortality vs kappa in England+Wales. (a) outbreak mortality vs time (b) mortality rate vs time. The jitter in this panel arises from the annual aging update step.

**Figure 6:**
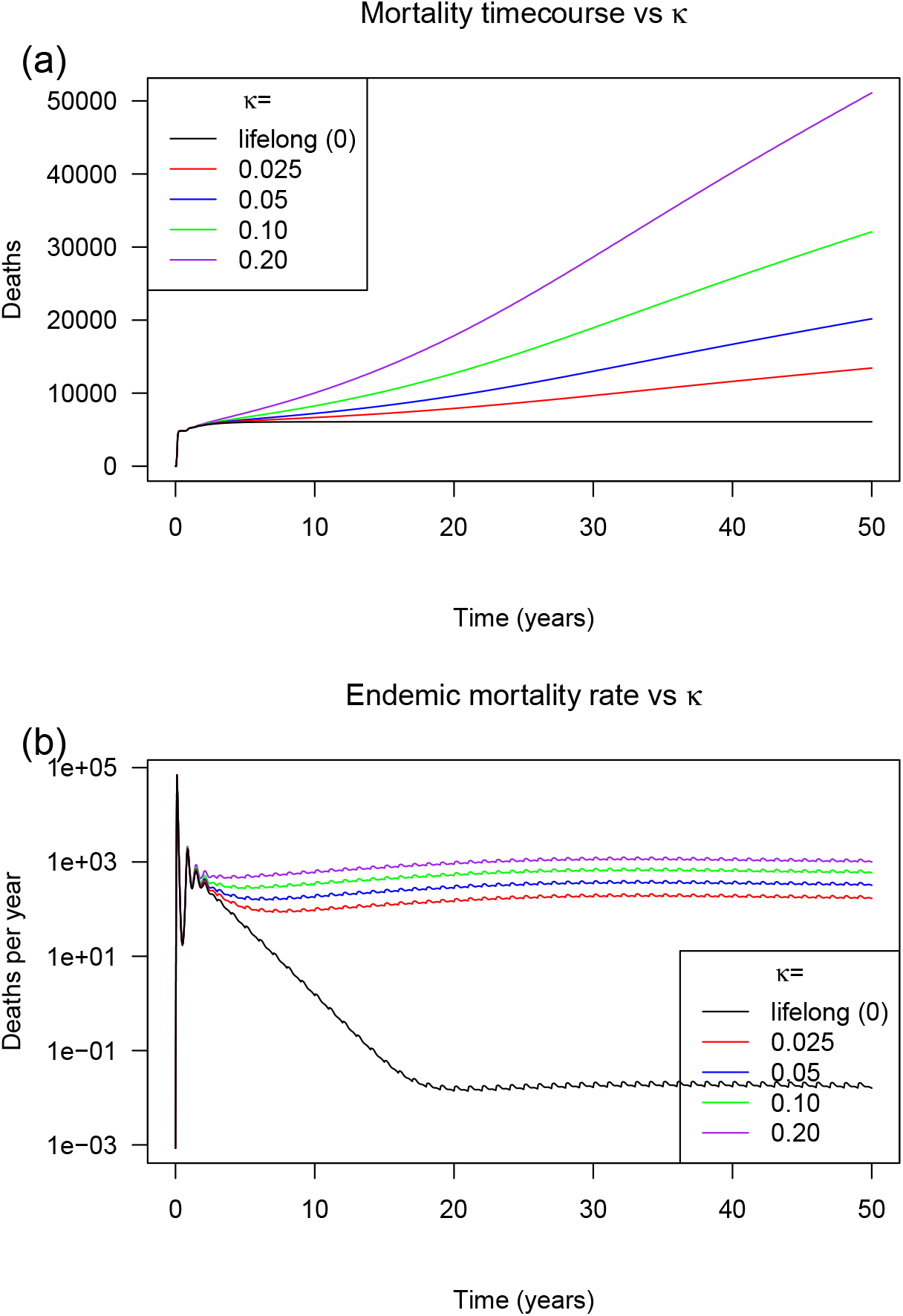
Mortality vs kappa in Singapore. (a) outbreak mortality vs time (b) mortality rate vs time. The jitter in this panel arises from the annual aging update step.

The initial death toll was large, irrespective of *κ*. When the 2-year death toll is used as a proxy for the common transitional mortality, estimates ranged between 7753 and 7996 for the England+Wales model. Equivalent figures of 5574 and 5767 were obtained for the Singapore model.

Without lifelong FII, the endemic mortality rate was rose rapidly with *κ* and was substantial at high *κ* (tables 5 and 6). At the highest value modelled (*κ* = 0.2), annual mortality was 864 and 1057 for the England+Wales and Singapore models respectively (tables 5 and 6), corresponding to 6.69% and 7.94% of total mortality respectively toward the end of the simulation. As a point of reference, influenza and pneumonia constitute 5% of deaths in England and Wales with an ongoing vaccination programme. However, it should be noted that other causes mortality rose from around 8400 to nearly 13000 as the population aged over the 50 year simulation period but since both the mortality from modelled outbreak and influenza/pneumonia arises from the same elderly population, this comparison may still be valid. Whether the new outbreak will contribute additively to influenza deaths is beyond the scope of this work but a novel ECoV and influenza will draw many of their fatalities from the same pool of the elderly.

**Table 5:** Vaccination - E+W. Mortality rates were averaged over t=48-50. ‘>60 @start’: vaccinating over 60s at start. ‘>20 @start’: vaccinating over 20s at start. ‘>60 20%/yr’: vaccinating 20% of over 60s each year. ‘>60 40%/yr’: vaccinating 40% of over 60s each year.

| $\kappa =$ | lifelong | 0.025 | 0.05 | 0.10 | 0.20 |
| --- | --- | --- | --- | --- | --- |
| Deaths after 2 years | 7753 | 7785 | 7817 | 7879 | 7996 |
| Endemic deaths (/yr) | 0 | 142 | 272 | 500 | 864 |
| Other causes (/yr) | 12919 | 12778 | 12649 | 12421 | 12059 |
| % of annual deaths | 0.00 | 1.10 | 2.10 | 3.87 | 6.69 |
| >60 @start - deaths avoided | 4036 | 3974 | 3917 | 3817 | 3657 |
| >20 @start - deaths avoided | 4292 | 4248 | 4207 | 4135 | 4020 |
| >60 20%/yr - Endemic deaths (/yr) | 0 | 100 | 192 | 360 | 637 |
| >60 20%/yr - % endemic deaths avoided | NA | 29.8 | 29.2 | 28.1 | 26.3 |
| >60 40%/yr - Endemic deaths (/yr) | 0 | 72 | 141 | 266 | 480 |
| >60 40%/yr - % endemic deaths avoided | NA | 49.0 | 48.3 | 46.9 | 44.4 |

**Table 6:**
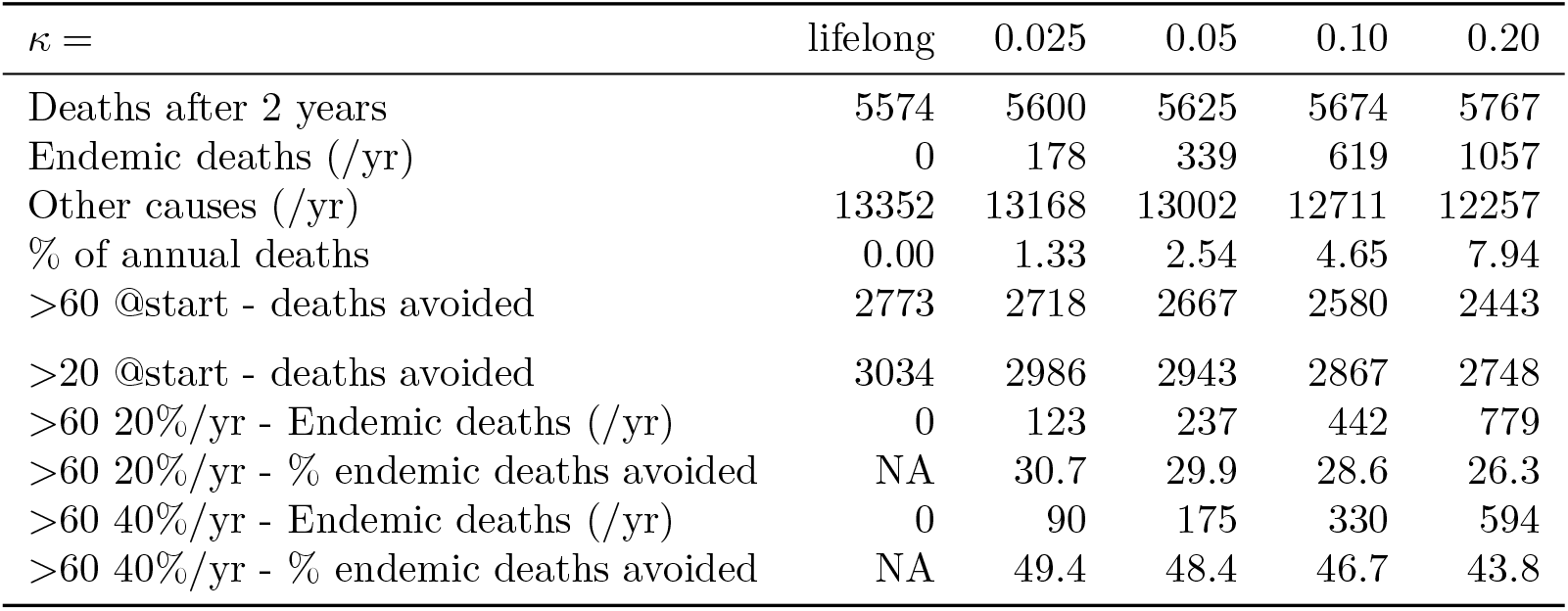
Vaccination - SG. Mortality rates were averaged over t=48-50. ‘>60 @start’: vaccinating over 60s at start. ‘>20 @start’: vaccinating over 20s at start. ‘>60 20%/yr’: vaccinating 20% of over 60s each year. ‘>60 40%/yr’: vaccinating 40% of over 60s each year.

| $\kappa =$ | lifelong | 0.025 | 0.05 | 0.10 | 0.20 |
| --- | --- | --- | --- | --- | --- |
| Deaths after 2 years | 5574 | 5600 | 5625 | 5674 | 5767 |
| Endemic deaths (/yr) | 0 | 178 | 339 | 619 | 1057 |
| Other causes (/yr) | 13352 | 13168 | 13002 | 12711 | 12257 |
| % of annual deaths | 0.00 | 1.33 | 2.54 | 4.65 | 7.94 |
| >60 @start - deaths avoided | 2773 | 2718 | 2667 | 2580 | 2443 |
| >20 @start - deaths avoided | 3034 | 2986 | 2943 | 2867 | 2748 |
| >60 20%/yr - Endemic deaths (/yr) | 0 | 123 | 237 | 442 | 779 |
| >60 20%/yr - % endemic deaths avoided | NA | 30.7 | 29.9 | 28.6 | 26.3 |
| >60 40%/yr - Endemic deaths (/yr) | 0 | 90 | 175 | 330 | 594 |
| >60 40%/yr - % endemic deaths avoided | NA | 49.4 | 48.4 | 46.7 | 43.8 |

### Vaccination

The use of vaccination to reduce the death toll during the the transition to endemicity was studied. It was reasoned that accelerating the establishment of FII by a single vaccination could reduce mortality during the transition to endemicity even if no subsequent vaccinations were administered since circulating infection would maintain protection thereafter. This was modelled by moving half of the initial population above either the age of 20 or 60 from the S state to the R state and examining outcomes with either lifelong FII or with 0.025 ≤ *κ* ≤ 0.2. The difference between the transition mortality in the base case and that of the intervention case is then an estimate of the lives saved by that intervention. The results can be seen in Tables 5 and 6.

With lifelong FII, vaccinating 50% of over-60s reduced deaths by 47.5% and 47% in the England+Wales and Singapore models respectively. (tables 1, 2, 5 and 6). The small shortfall arose from under-60s being fatally infected for the first time as they aged as is apparent when deaths with the over-60s vaccination is compared against that from vaccinating over-20s. That comparison also demonstrates the diminishing returns from expanding the scope of vaccination to younger ages as a means of reducing transitional mortality.

With shorter FII half-lives, the control of endemic immunity becomes increasingly important but requires periodic vaccination programmes. A crude attempt was made to model periodic vaccination by moving a fraction of the living population each year to the R state. Two rates were examined, a effective vaccination rate of 20% and 40% of the over-60s population per year and the impact of these interventions on endemic mortality was examined (tables 5 and 6). As expected, the protection improved with vaccination rate and fell when the FII state was shorter-lived (i.e. higher *κ*). The vaccination gains appear poor: vaccinating 40% of the >60 year olds annually would be expected to yield much more than a 40% gain when FII is longer-lived. This is a consequence of the vaccination being inefficiently targeted at many that were still immune. Vaccination on a periodic basis would have targeted those more likely to be losing FII but ODE modelling is ill-suited to this. Better analysis of vaccination programme details will require the use of other modelling approaches.

## Discussion

This study proposes that ECoV and more recent pathogenic coronaviruses share a set of immunological properties that have not been fully observed in either group of viruses. This proposal can be modelled to generate falsifiable predictions of outbreak behaviour.

A central feature of this proposal is that immunity acquired safely when young, is maintained throughout life by reinfection, protects the subject from death when they reach the age when they would otherwise be vulnerable. To this end, an ODE model has been constructed to model long-term evolution of ECoV type outbreaks incorporating aging of the population. Contact matrices incorporate key interactions between the different age groups.

Simulations using this model showed two components to virus mortality. The first component occurs between the start of the outbreak and the establishment of an endemic steady state. Therafter, the population suffers from the ongoing endemic mortality rate. Decreasing durability of FII shifts the mortality away from a initial quantum towards an ongoing death toll.

The immediate utility of this proposal lies in the predictions that emerge from this model if it should be applicable to Covid-19. That can only be known if the epidemiology of reinfection of SARS-2 is better understood. Indeed, elucidating that is crucial to the management of Covid-19. With regard to this proposal, the mortality and morbidity experienced in reinfection will determine whether FII exists. The model in its current simple form postulates that the FII state is fully infectious and this may also need to be modified in the light of experience. The best course of action remains the deployment of a vaccine if one is available irrespective of whether this proposal holds true. If subsequent data shows an infectible FII state exists, then considerations derived from this model may come into play and these are now discussed.

If the FII state is sufficiently long-lived and has an acceptable level of morbidity on reinfection, there is the option of letting the outbreak evolve towards endemicity. The role of the vaccine is the suppression of transition mortality and assuming that the vaccine used establishes an immune state similar to that of FII, all this would require is a single application of the vaccination programme covering the entire population. Circulating infection will immunize neonates and maintain FII thereafter.

If the FII state is shorter-lived, continued vaccination will rest on whether the endemic mortality rate and morbidity of the endemic state are acceptable. A programme targeted at the elderly and those with co-morbidities only may be adequate.

The properties of the vaccine also matter. If a vaccine conferred an unexpectedly long immunity against reinfection, then that vaccine could establish herd immunity and also be the basis of an global eradication effort. However, few vaccination programmes achieve total coverage and if this route is taken, significant fractions of the population may be unvaccinated and they remain vulnerable to fatal infection if vaccination is discontinued and the virus reintroduced from elsewhere. If immunity elicited by this supra-natural vaccine decays to the V state, the programme could still be discontinued at a later stage and circulating infection restablished to protect the population. However, circulating infection must be established before immunity wanes further to put the population back to the immunologically naive and fatally vulnerable S state.

## Materials and Methods

### Software

Simulations were performed using R 3.6.1 and RStudio 1.3.1093 (R Core Team, 2019; RStudio Team, 2016).

### States

States are consists of a vector of 180 elements, 90 for each sex. The elements represent counts for that sex in 1 year wide age groups up to the age of 90 years.

To avoid clutter, the sex indices have been omitted in the equations shown below.

### Demographic data

England and Wales mortality data for 2019 was obtained from NOMIS, downloaded on 19/10/2020. England and Wales population structure for 2019 was obtained from NOMIS, downloaded on 19/10/2020. NOMIS is at https://www.nomisweb.co.uk/

Singapore mortality data for 2018 was obtained from Singstat. Singapore population structure for 2018 was obtained from Singstat. Singstat is at https://www.singstat.gov.sg/

### IFR data

Covid-19 IFR data was obtained from Ghisolfi *et al*, 2020. However, the 10 year wide bands it was provided in resulted in discontinuous jumps in the age profiles of many metrics. Also, the death rate changes very significantly across that 10 year band. This issue was addressed by fitting an exponential relationship to data form ages above 50 as the IFR takes very small values below that and the quality of fit there is inconsequential to the results of simulations.

A final issue is that the final age bin for states is for age ≥ 89 and an FOI should be specified on the same basis. In this simulation, the effective age of that final bin was arbitrarily specified as 95 to account for that.

### Contact matrix

Contact matrices were obtained from Prem *et al*, 2017.

The contact matrices are defined with ages grouped into 5 year bins spanning 0-80 years of age. Also, they are not sex-specific. The contract matrices were block-expanded and reweighted to cover the 0-90 age range used in this simulation.

It was assumed that contact matrices did not change significantly over the duration of the simulation.

### Model equations

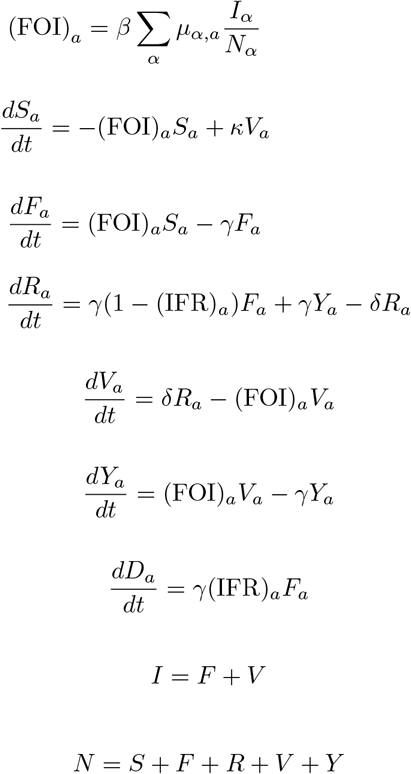

### Aging

Aging for the S,F,R and Y states was implemented by annually shifting the contents of age bins in the state vectors up one year. The final age ≥ 79 bin is not shifted but a lso accumulates t he contents of the the age 78 bin at each update.

A consequence of stepped aging is periodic jitter in model parameters. Some rate variables were averaged over 40 periods (2 years) in tables to reduce the influence of this.

### Calibration of beta

The contact matrix plays a central role in determining the dynamics of the model since it describes the potentially infectious contacts between age groups. It has been suggested that there is an age-dependent susceptibility to infection and the contact matrix was modified to reflect the reduced susceptibilities amongst the young as proposed in Davies *et al*, 2020.

When the simulations for different populations are to be compared, a decision has to be made on what is to be held invariant between the populations. In this case, either *R*_0_ or *β* could be fixed. There did not appear to be strong grounds for holding *R*_0_ constant since the social connectedness of populations, and therefore *R*_0_, can be expected to differ. *β* is associated with the probability of infection for a given contact rate and may perhaps be more of an intrinsic feature than *R*_0_ although cultural practices may be expected to influence the risk of infection on social contact too. A decision was taken to derive the *β* required to achieve *R*_0_ = 2.5 in the England+Wales simulation and use the same value of *β* for the Singapore simulation.

The relationship between *β* and *R*_0_ was established by running simple SIR simulations using a specified contact matrix over a range of *β* values and calculating the associated *R*_0_ from the asymptotic terminal fraction infected at herd immunity using the final size equation (Heffernan, Smith and Wahl, 2005). The required *β* of 93.5 for *R*_0_ = 2.5 in the England+Wales simulation was then obtained by interpolation. This was equivalent to *R*_0_ being 2.94 in the Singapore case. The raised *R*_0_ in the latter reflected t he higher connectedness of the Singapore population as embedded in the weights of the contact matrix.

## Supporting information

Zip archive containing RMarkdown file that generated this manuscript, data and simulation code.

## Data Availability

All data and code used to generate this manuscript are deposited in supplementary material.

## Supplementary information

The RMarkdown script that ran all simulations and generated this manuscript is deposited as supplementary data.

## Acknowledgments

This study was entirely self-funded.

The use of bibliographic services *via* the University of Cambridge is gratefully acknowledged.

## Notes

### Competing Interest Statement

The authors have declared no competing interest.

### Funding Statement

This work was entirely self-funded.

### Summary of Updates

Errors fixed in calibrating beta and in one of the tables. Model R0 changed from 1.8 to 2.5 which is used more frequently in other modelling manuscripts. Text adjusted to reflect changes.

