## Supplementary material for "A model of endemic coronavirus infections": Zip archive containing RMarkdown file that generated this manuscript, data and simulation code.: data.R

```
# base data
# SG mortality data (SG, 2018,https://www.tablebuilder.singstat.gov.sg/publicfacing/createDataTable.action?refId=13249)
# These are age-specific death rates in per 1000.
# Each has missing value so I have filled in with nearby value.
# To keep lengths consistent, I have filled 89-90 with 90+ value.
sg.mortality.table.mel<-c(2.3,0.1,0.1,0.1,0.1,
                       rep(c(0.1,0.1,0.3,0.4,0.3,0.5,0.6,0.9,1.8,2.8,5.0,8.2,12.5,22.4,35.9,62.8),each=5),
                       102.9,102.9,102.9,102.9,169.3
)/1000;
sg.mortality.table.fem<-c(1.9,0.1,0.1,0.1,0.1,
                       rep(c(0.1,0.1,0.1,0.2,0.2,0.3,0.4,0.5,1.0,1.8,2.7,4.4,6.6,11.3,22.3,43.2),each=5),
                       74.1,74.1,74.1,74.1,149.1
)/1000;
# These are CDR in % of population.  From NOMIS, 2019.
ew.mortality.table.mel<-c(0.426,0.013,0.013,0.013,0.013,
                          rep(c(0.007,0.010,0.027,0.051,0.062,0.081,0.114,0.160,0.255,0.380,0.563,0.880,1.408,2.178,3.738,6.616),each=5),
                          11.915,11.915,11.915,11.915,22.424)/100;

ew.mortality.table.fem<-c(0.352,0.014,0.014,0.014,0.014,
                          rep(c(0.007,0.007,0.016,0.021,0.028,0.044,0.067,0.103,0.164,0.244,0.367,0.584,0.902,1.471,2.592,4.829),each=5),
                          9.233,9.233,9.233,9.233,20.033)/100;
# These are from https://apps.who.int/gho/data/view.main.60850, 2016
# Original figures in fraction rather than %, converted for here by x100.
#ke.mortality.table.mel<-c(4.1,0.4,0.4,0.4,0.4,
#                          rep(c(0.2,0.1,0.2,0.3,0.4,0.4,0.6,0.7,0.8,1.1,1.3,2.0,3.0,4.7,7.5,12.2),each=5),
#                          22.6,22.6,22.6,22.6,22.6)/100;

#ke.mortality.table.fem<-c(3.4,0.3,0.3,0.3,0.3,
#                          rep(c(0.1,0.1,0.1,0.2,0.3,0.3,0.4,0.5,0.6,0.7,0.9,1.4,2.3,3.8,6.4,10.8),each=5),
#                          20.5,20.5,20.5,20.5,20.5)/100;


# Age distribution
# Note this is not in equilibrium with mortality data.
# SG is still working through consequences of increased life expectancy.
# These are 2018 figures.
# Note the 90+ band started 1 year early because of top element in age data structure is 89+.
# https://www.tablebuilder.singstat.gov.sg/publicfacing/createDataTable.action?refId=15689
sg.age.table.mel<-c(rep(floor(c(88607,90485,92319,106330,122575,129837,110705,111745,111210,120509,132636,139669,127127,98644,60956,39346,22952)/5),each=5),
                    rep(floor(11429/5),4),floor(11429/5)+5107);
sg.age.table.fem<-c(rep(floor(c(84014,85311,87903,99393,114471,121121,107169,112880,116346,126820,135209,140727,130421,104261,69099,50190,32280)/5),each=5),
                    rep(floor(19814/5),4),floor(19814/5)+12319);
# From NOMIS, 2019 data.
ew.age.table.mel<-c(rep(floor(c(1775185,1907778,1813263,1675109,1901759,2047905,2005133,1946773,1783713,1938616,2034595,1916618,1622414,1442704,1416997,960727,675179)/5),each=5),
                    rep(floor(371814/5),4),floor(371814/5)+141605+30280+1966+101);
ew.age.table.fem<-c(rep(floor(c(1687981,1817840,1725110,1585362,1791131,1973985,2005453,1979177,1802515,1980077,2094159,1973812,1685662,1539329,1543023,1107998,855376)/5),each=5),
                    rep(floor(561094/5),4),floor(561094/5)+272528+86433+9211+612);
# From https://population.un.org/wpp/Download/Standard/Population/, 2020 estimates
#ke.age.table.mel<-c(rep(floor(c(3558000,3508000,3395000,3022000,2619000,2167000,1993000,1704000,1362000,1055000,782000,574000,411000,279000,161000,76000,37000)/5),each=5),
#                    rep(floor(12000/5),4),floor(12000/5)+2+0+0);
#ke.age.table.fem<-c(rep(floor(c(3486000,3461000,3341000,2989000,2617000,2191000,2030000,1743000,1397000,1088000,822000,628000,477000,351000,220000,117000,65000)/5),each=5),
#                    rep(floor(25000/5),4),floor(25000/5)+4+0+0);


# IFR distribution (HIC) https://gh.bmj.com/content/bmjgh/5/9/e003094.full.pdf
# no: no comorbidities; co: with comorbidities.
# Using the 10 year wide age groups causes very discontinuous simulation profiles vs age
# as the IFR changes very rapidly with age.
# Noting that the fast changes occur above 50, I will fit data for over-50s age groups only.
# The error when projected back to younger age groups is inconsequential as IFR values
# are so low.
group.offset<-7; # how far into age group to place age for fitting.
ifr.mel.no.tab<-data.frame(age=group.offset+c(50,60,70,80),ifr=0.01*c(0.0185,0.1105,0.4755,3.7759));
ifr.fem.no.tab<-data.frame(age=group.offset+c(50,60,70,80),ifr=0.01*c(0.0112,0.0438,0.1749,1.1708));
ifr.mel.co.tab<-data.frame(age=group.offset+c(50,60,70,80),ifr=0.01*c(0.7865,2.008,4.3483,19.9581));
ifr.fem.co.tab<-data.frame(age=group.offset+c(50,60,70,80),ifr=0.01*c(0.6353,1.2395,2.3906,7.0568));

gen.ifr.fit<-function(ifr.data) {
  lm.fit<-lm(log(ifr)~age,data=ifr.data);
  exp(lm.fit$coefficients[1]+lm.fit$coefficients[2]*c(1:89,95));
}

ifr.mel.no<-gen.ifr.fit(ifr.mel.no.tab);
ifr.fem.no<-gen.ifr.fit(ifr.fem.no.tab);

ifr.mel.co<-gen.ifr.fit(ifr.mel.co.tab);
ifr.fem.co<-gen.ifr.fit(ifr.fem.co.tab);

# Comorbidity vs age
# A crude fit to supp figure data in above paper.
# Actually prepared from HIC country so validity can be questioned elsewhere.
no.comorb.fem<-approx(c(0,10,50,90),c(0.98,0.96,0.7,0.1),seq(0.5,89.5,1))$y;
no.comorb.mel<-approx(c(0,10,50,90),c(0.98,0.96,0.65,0.1),seq(0.5,89.5,1))$y;

# Read in a contact matrix
# rows == individual
# cols == their contacts
# Check by old individual has young contacts but young individual doesn't have old contacts.
sg.alllocs<-as.matrix(read.csv("SG_all_locations_contacts.csv",header=F));
uk.alllocs<-as.matrix(read.csv("UK_all_locations_contacts.csv",header=F));
#ke.alllocs<-as.matrix(read.csv("~/projects/sisccov/KE_all_locations_contacts.csv",header=F));

# Infection susceptibility factors.
# From Davies et al, Nature Medicine.  Extracted from graph.
davies.sigma<-c(rep(0.4,10),rep(0.38,10),rep(0.79,10),rep(0.86,10),rep(0.80,10),rep(0.82,10),rep(0.88,10),rep(0.74,20));
# From Ayoub et al, 2020.  medrxiv
ayoub.sigma<-c(rep(0.05,10),rep(0.06,10),rep(0.34,10),rep(0.57,10),rep(0.69,10),rep(0.79,10),rep(1,10),rep(0.94,10),rep(0.88,10));

# Create objects to make demographics more manageable.
ew<-list(long.label="England+Wales",
         short.label="E+W",
         age.table.mel=ew.age.table.mel,
         age.table.fem=ew.age.table.fem,
         mortality.table.mel=ew.mortality.table.mel,
         mortality.table.fem=ew.mortality.table.fem,
         contactMatrix=uk.alllocs);
sg<-list(long.label="Singapore",
         short.label="SG",
         age.table.mel=sg.age.table.mel,
         age.table.fem=sg.age.table.fem,
         mortality.table.mel=sg.mortality.table.mel,
         mortality.table.fem=sg.mortality.table.fem,
         contactMatrix=sg.alllocs);
```
